# Safety, Tolerability and Efficacy of 40Hz Sensory Stimulation for Alzheimer’s Disease

**DOI:** 10.1101/2023.03.23.23287637

**Authors:** Mihály Hajós, Alyssa Boasso, Evan Hempel, Monika Spokayte, Alex Konisky, Chandran V. Seshagiri, Vitella Fomenko, Kim Kwan, Jessie Nicodemus-Johnson, Suzanne Hendrix, Brent Vaughan, Jonathan T. Megerian, Zach Malchano

## Abstract

Alzheimer’s Disease (AD) is a multifactorial, progressive neurodegenerative disease that disrupts cognitive function through maladaptive misfolded proteins, abnormal neuroimmune responses, and disordered neuronal network activities. Despite continued scientific advances in the understanding of AD biology, there remains an unmet need for safe and effective disease-modifying treatments. Sensory stimulation is an emerging therapeutic approach which has demonstrated disease-modifying effects in preclinical transgenic models of AD. This randomized, sham-controlled, clinical trial (OVERTURE; NCT03556280) evaluated the feasibility and safety of 40Hz auditory and visual stimulation with the CogTx-001 medical device in 70 participants with mild to moderate AD, administered as daily, 1-hour active stimulation (as compared to sham stimulation) over a 6-month period. Primary endpoints of the therapy showed that it was well-tolerated, showed high adherence and demonstrated a favorable safety profile. Secondary outcomes included exploratory outcomes measures such as ADCS-ADL and MMSE scores, which demonstrated significant effects on functional and cognitive abilities. Additionally, sensory stimulation also showed a significant reduction in brain volume loss and cortical thinning, without changes in amyloid PET signal in active versus sham groups. These encouraging results justify further development of 40Hz sensory stimulation as a safe and potentially disease-modifying therapy for AD patients.

## INTRODUCTION

Neuronal network dysfunctions, including abnormal oscillatory rhythmic activity and network hypersynchrony, widely overlap with brain regions that develop pathological hallmarks and atrophy in Alzheimer’s disease (AD) patients^1–3^. Modulation and restoration of these functional network alterations may provide new strategies for disease-modifying treatments in AD. Cognito Therapeutics, Inc. has developed an innovative AD medical device therapy that uses a non-invasive sensory stimulation system for at-home use. Safety, tolerability and early potential effects on biomarkers of 40Hz sensory stimulation have been previously evaluated by two clinical trials, with 2-3 months of treatment^4, 5^. However, in this randomized, sham (placebo)-controlled (OVERTURE; NCT03556280) clinical trial the safety, tolerability, and clinical efficacy of Cognito Therapeutics CogTx-001 medical device were evaluated by providing daily, 1-hour 40Hz auditory and visual stimulation to participants with mild to moderate AD over a 6-month period. During the screening phase, stimulation intensities were tailored to each participant in a clinical setting and the presence of treatment-evoked 40Hz steady-state oscillations was verified for study inclusion.

Accumulating experimental evidence suggests that evoked 40Hz gamma oscillation has the potential to alleviate AD pathology and preserve cognitive function^6^. Sustained activation of synchronized 40Hz gamma oscillations via optogenetic or sensory stimulation in AD transgenic mice was shown to effectively reduce AD pathology and slow disease progression^7–13^. These results demonstrated that neuronal 40Hz steady-state oscillations, evoked by daily visual and auditory stimulation over several weeks was accompanied by multiple downstream biological effects, including attenuation of synaptic loss and neurodegeneration, reduction in amyloid and tau pathologies, and improvement in circadian rhythm as well as learning and memory in various AD mice models^6, 13^. In the Overture trial, a broad range of established AD scales measuring cognition, function, and biomarkers were included in the design of the study to measure treatment effects. Potential changes in cognition and function were evaluated by a number of clinical instruments, including the Mini-Mental State Exam (MMSE), Alzheimer’s Disease Cooperative Study Activities–Cognitive Subscale (ADAS-Cog14), Alzheimer’s Disease Cooperative Study Activities of Daily Living (ADCS-ADL), Clinical Dementia Rating (CDR), and the AD Composite Score (ADCOMS, and its version, MADCOMS tailored for mild to moderate AD participants). As an exploratory trial, without previous studies addressing clinical benefits of this treatment in participants, all efficacy outcome measures were considered exploratory in this trial. In addition to assessing cognitive and functional outcomes, the trial measured changes in amyloid pathology and brain atrophy by volumetric MRI measurements. Results of the OVERTURE trial support the initiation of a randomized, controlled pivotal clinical trial to further explore potential clinical benefits of 40 Hz sensory stimulation in AD patients.

## RESULTS

### SAFETY AND TOLERABILITY

In this multicentered clinical trial, 135 participants were screened, 76 participants were randomized 2:1 to active vs sham (placebo) arm (**Fig. 1**), and 74 participants received treatment; 46 in the active arm and 28 in the sham arm. The trial was completed by 33 (71.74%) participants who used active stimulation and 20 (71.43%) participants who used sham stimulation with a 28.38% overall dropout rate. Participants in the active arm received 40Hz auditory-visual sensory stimulation that effectively evoked 40Hz steady-state oscillations, whereas in the sham arm sensory stimulation was designed to be ineffective in evoking 40 Hz steady-state oscillations. Participants in both arms used the medical device at home daily for one hour and followed the same treatment protocol over a 6-month period. Participants, their care partners, and raters at the clinical sites were blinded to treatment allocation. Trial inclusion and exclusion criteria are listed in **Extended Data Table S1**. Demographic and baseline characteristics of sham and active arm participants are presented in **Table 1** and were generally balanced across treatment groups, with the sham group showing lower MMSE and higher CDR-SB scores at baseline. APOE4 status, amyloid PET SUVR, and whole brain volumes were comparable in both treatment groups at baseline.

**Figure 1:**
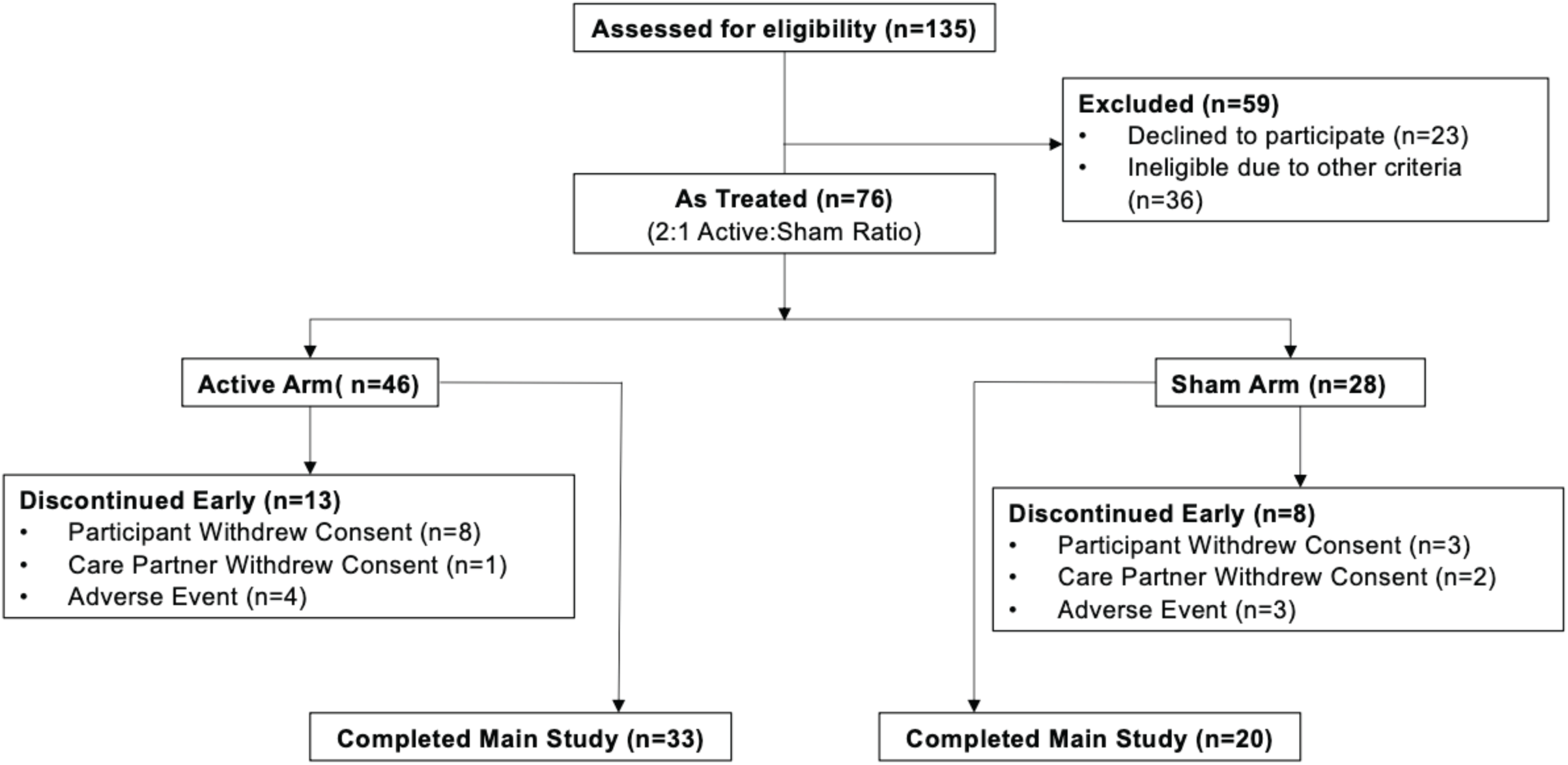
CONSORT Chart. Diagram depicts participant allocation to treatment arm in the safety population. Of the 76 participants enrolled, 2 did not receive treatment and are not included in the safety population. One participant that was randomly assigned to the sham group received active therapy during the trial, therefore, is included in the as-treated safety population (active: n=46; sham: n=28). Four additional participants in the active group withdrew from the study without a follow-up assessment and therefore are not included in the efficacy analysis (i.e., ITT as treated population, active n=42, sham n=28). Details are provided in the methods. Inclusion and exclusion criteria can be found in the supplementary appendix (**Extended Data Table 1**).

**Table 1:** Baseline Demographics for the ITT As Treated Population

|  | ITT As Treated |  |
| --- | --- | --- |
|  | Active (n=43) | Sham (n=27) |
| **Sex** |  |  |
| Female No (%) | 24 (55.8%) | 12 (44.4%) |
| Male No (%) | 19 (44.2%) | 15 (55.6%) |
| **Age, mean (SD)** |  |  |
|  | 69.7 (7.99) | 75.6 (10.04) |
| **Race** |  |  |
| Other | 1 (2.3%) | 0 (0.0%) |
| White | 42 (97.7%) | 27 (100%) |
| **APOE4** |  |  |
| Heterozygous | 16 (37.2%) | 10 (37.0%) |
| Homozygous | 3 (7.0%) | 2 (7.4%) |
| Non-carrier | 20 (46.5%) | 15 (55.6%) |
| Carriers* | 3 (7.0%) | 0 (0.0%) |
| Unknown | 1 (2.3%) | 0 (0.0%) |
| **Florbetapir SUVr\*\*** |  |  |
| N | 43 | 27 |
| Mean (SD) | 1.3 (0.25) | 1.3 (0.25) |
| Amyloid + | 32 | 18 |
| Amyloid - | 11 | 9 |
| **ADCS-ADL (Total)** |  |  |
| N | 37 | 26 |
| Mean (SD) | 66.9 (8.54) | 63.3 (10.65) |
| **ADCS-ADL (Instrumental)** |  |  |
| N | 43 | 26 |
| Mean (SD) | 44.2 (8.35) | 42.0 (9.91) |
| **ADCS-ADL (Basic)** |  |  |
| N | 43 | 27 |
| Mean (SD) | 21.4 (1.36) | 21.2 (1.28) |
| **MMSE** |  |  |
| N | 43 | 27 |
| Mean (SD) | 21.4 (3.28) | 19.7 (3.85) |
| **CDR-Sum of Boxes** |  |  |
| N | 43 | 27 |
| Mean (SD) | 4.6 (1.54) | 6.2 (2.79) |
| **MADCOMS** |  |  |
| N | 43 | 26 |
| Mean (SD) | 7.0 (1.96) | 7.6 (2.63) |
| **Whole Brain Volume** |  |  |
| N | 36 | 20 |
| Mean (SD) | 1060.7 (129.35) | 1017.7 (122.80) |

Tolerability of the treatment was assessed via device-recorded usage throughout the treatment period. High adherence to daily device therapy was calculated as the percentage of study days when the device was used for 45 minutes or more. Average adherence across all participants who completed the main study was 85.1%. Among completers, participants who used the sham device had a slightly higher average adherence rate (active: 81.3%, sham: 92.1%, p = 0.014, Wilcoxon Rank Sum test). Rate of early terminators was the same (28%) in both active and sham groups and showed similar adherence rates (active: 77%, sham:74%).

The safety population included all trial participants who received at least one treatment (n=46 active and n=28 sham). There were no deaths reported during the study. No unanticipated adverse events (AEs) related to device use over a 6-month treatment period were reported. Treatment-emerging AEs (TEAEs) and treatment-related AEs (TRAEs) are shown in **Table 2**. TEAEs were reported by 30 (65%) participants in the active group and 20 (71%) participants in the sham group. TRAEs were reported in 19 (41%) participants in the active group and 8 (29%) participants in the sham group. Serious AEs (SAEs) were reported by 3 (6.5%) participants in the active group and 3 (10.7%) participants in the sham group. Among the TRAEs (defined as possibly, probably, or definitely related to treatment/device by the site principal investigator), all were mild except for 3 events; 2 moderately severe events (tinnitus, active participant; agitation, sham participant) and 1 severe event (transient confusional state, active participant occurring 39 days after starting treatment and resolved completely). Tinnitus was reported by 7 (15%) participants in the active arm and none in the sham arm, headache reported by 10 participant (22%) in the active and 3 (11%) participants in the sham groups. Other TRAEs reported in the sham group occurred at low incidence (2 participants, 7%) rate and included 2 participants eye pain and 2 participants with disorientation. The Columbia–Suicide Severity Rating Scale, administered throughout the study, did not suggest a substantial increase in suicidality among participants in the active arm. MRI image analysis, reviewed by a neuroradiologist blinded to treatment arms at a central reading facility (Biospective Imaging Corp), demonstrated neither vasogenic edema and sulcal effusions (ARIA-E) nor hemosiderin deposits (ARIA-H) greater than 10 mm in any participants who completed the trial (n = 53) and who also had 6-month magnetic resonance imaging (n = 52). In summary, the data supports the safety and tolerability of Cognito’s medical device in mild to moderate AD participants.

**Table 2:** Treatment-Emerging Adverse events (TEAEs) and Treatment-Related Adverse Events (TRAEs) as Safety Population

| Treatment Emergent Adverse Events (TEAE) | Active | Sham | Treatment Related Adverse Events (TREA) | Active | Sham |
| --- | --- | --- | --- | --- | --- |
| System Order Class<br>Preferred Term | N=46<br>n (%) | N=28<br>n (%) | System Order Class | N=46<br>n (%) | N=28<br>n (%) |
| Number of Subjects with any TEAE | 30 (65.2) | 20 (71.4) | HeadAche | 10 (21.7) | 3 (10.7) |
| Ear and labyrinth disorders | 9 (19.6) | 0 (0.0) | Tinnitus | 7 (15.2) | 0 (0.0) |
| Tinnitus | 7 (15.2) | 0 (0.0) | Eye Pain | 0 (0.0) | 2 (7.1) |
| Eye disorders | 3 (6.5) | 3 (10.7) | Disorientation | 0 (0.0) | 2 (7.1) |
| Eye pain | 0 (0.0) | 2 (7.1) |  |  |  |
| Gastrointestinal disorders | 3 (6.5) | 3 (10.7) |  |  |  |
| General disorders and administration site conditions | 2 (4.3) | 4 (14.3) |  |  |  |
| Oedema | 0 (0.0) | 2 (7.1) |  |  |  |
| Infections and infestations | 7 (15.2) | 7 (25.0) |  |  |  |
| Nasopharyngitis | 0 (0.0) | 6 (21.4) |  |  |  |
| Injury, poisoning and procedural complications | 2 (4.3) | 2 (7.1) |  |  |  |
| Skin abrasion | 0 (0.0) | 4 (14.3) |  |  |  |
| Investigations | 3 (6.5) | 0 (0.0) |  |  |  |
| Musculoskeletal and connective tissue disorders | 6 (13.0) | 1 (3.6) |  |  |  |
| Nervous system disorders | 14 (30.4) | 10 (35.7) |  |  |  |
| Headache | 10 (21.7) | 4 (14.3) |  |  |  |
| Agitation | 2 (4.3) | 2 (7.1) |  |  |  |
| Dizziness | 2 (4.3) | 2 (7.1) |  |  |  |
| Disorientation | 0 (0.0) | 2 (7.1) |  |  |  |
| Psychiatric disorders | 7 (15.2) | 4 (14.3) |  |  |  |
| Anxiety | 1 (2.2) | 2 (7.1) |  |  |  |
| Restlessness | 0 (0.0) | 2 (7.1) |  |  |  |
| Respiratory, thoracic and mediastinal disorders | 1 (2.2) | 2 (7.1) |  |  |  |
| Skin and subcutaneous tissue disorders | 4 (8.7) | 0 (0.0) |  |  |  |

### CLINICAL EFFICACY MEASURES

For evaluating treatment efficacy, a broad range of efficacy outcomes were selected based on their demonstrated utility to evaluate potential disease-modifying treatment effects in AD clinical trials^14, 15^. Prespecified primary outcome measures included an optimized AD Composite Score (ADCOMS) measure for improved sensitivity towards mild and moderate AD patients (MADCOMS), which consisted of 7 items: 3 ADAS–Cog subscale items, 2 MMSE items, and 3 CDR-SB items. MADCOMS scores in sham arm participants showed a worsening change from 7.6±2.63 at baseline to 9.2±4.28 at 6-month, and active arm participants showed a change from 7.0±1.96 at baseline to 7.9 ±2.98 at 6-month, without significant difference between arms (p=0.683) (**Fig. 2a**). Additional prespecified primary outcome measures were the Alzheimer’s Disease Cooperative Study Activities – Cognitive Subscale (ADAS-Cog14) and Clinical Dementia Rate Sum of Boxes (CDR-SB). The ADAS-Cog14 covers all cognitive areas in dementia and is particularly sensitive to changes in verbal memory and word recognition^16^. In the sham group participants, there was a 4.5-point change in ADAS-Cog14 scores (34.8 ±13.03 at baseline, and 39.3 ±17.76 at 6-months), indicating worsening, with a similar, 2.8-point change in active arm participants as well (baseline: 27.9±8.58, 6-month: 30.7±11.9) (p=0.4083) (**Fig. 2b**). The CDR-SB is designed to capture cognition and function, and has been used in multiple clinical trials as primary or secondary endpoint^17^. Participants in sham arm showed 1.0 points change in CDR-SB, from 6.2 ±2.79 at baseline to 7.2±3.72 at 6-months, and participants in active arm showed 0.7 points change in CDR-SB, from 4.6±1.54 at baseline to 5.3 ±2.16 at 6-months, without significant separation between arms (p=0.8941) (**Fig. 2c**).

**Figure 2:**
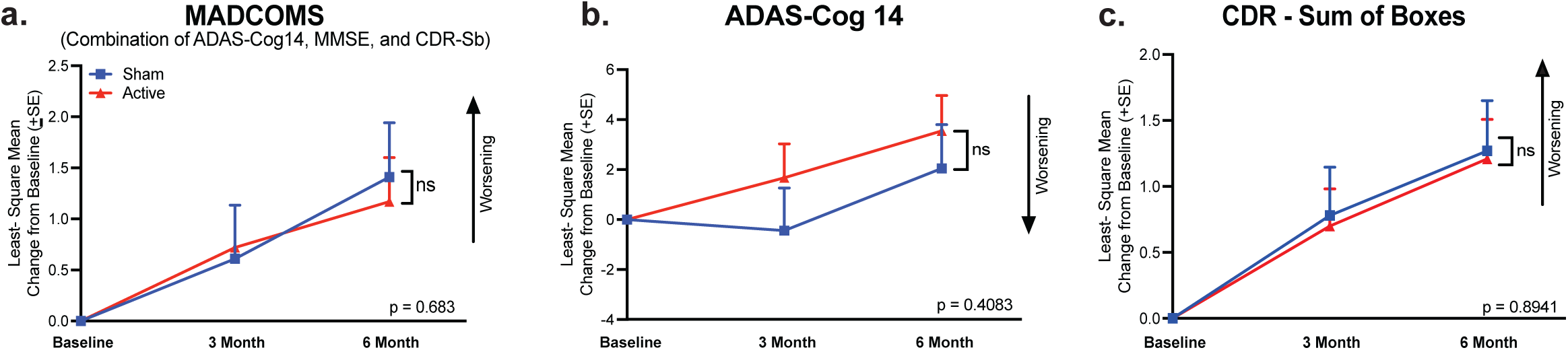
Primary Outcome Measures. Participants in the Overture trial did not show significant changes on traditional primary outcome measures. **a,** MADCOMS (combination of ADAS-Cog14, MMSE, and CDR-sb) change from baseline scores in sham (n=26) vs active (n=43) participants (p=0.683). **b,** ADAS-Cog 14 change from baseline scores in sham (n=26) vs active (n=43) participants (p=0.408). **c,** CDR – Sum of Boxes change from baseline scores in sham (n=27) vs active (n=43) in participants (p=0.894). All analyses were conducted using a linear mixed model with significance established as p<0.05. Arrows represent the direction of worsening.

A secondary outcome measure in this trial was the ADCS-ADL, which is a sensitive clinical instrument used to assess patients’ everyday function from basic abilities such as self-care to more complex tasks such as operating simple home devices and dealing with finances^18^. ADCS-ADL total scores showed a decline from 63.3±10.65 at baseline to 49.9±19.17 at 6-months in sham arm participants, whereas active arm participants maintained their total ADCS-ADL scores near baseline levels, showing a smaller decline from 66.9 ±8.54 at baseline to 63.5 ±9.01 at 6-months. The difference in decline in ADCS-ADL Total scores between sham and active arm participants was significant (ΔLSMean ± SE = 6.61 ± 1.84, p=0.0004) (**Fig. 3a**). Furthermore, both ADCS-ADL instrumental and basic scores showed a similar treatment response. ADCS-ADL Instrumental scores reflect more complex activities of daily living and more likely to detect early effects of cognitive decline, making it helpful in diagnosing early dementia. ADCS-ADL Instrumental scores declined from 42.0 ±9.91 at baseline to 32.0 ±14.16 at 6-months in sham arm participants, while the decline in active arm participants was less pronounced changing from 44.2±8.35 at baseline to 42.1±8.03 at 6-months. There was a significant difference (ΔLSMean ± SE = 5.40 ± 1.38; p =0.0001) in decline in ADCS-ADL instrumental scores between sham and active arms participants (**Fig. 3b**). ADCS-ADL Basic scores showed a decline from 21.2±1.28 at baseline to 17.9±5.80 at 6-months in sham arm participants, whereas, active arm participants scores declined from 21.4±1.36 at baseline to 21.2±1.51 at 6-months, and there was a significant difference in decline between sham and active arms participants (ΔLSMean ± SE = 1.62 ± 0.55, p=0.004) (**Fig. 3c**).

**Figure 3:**
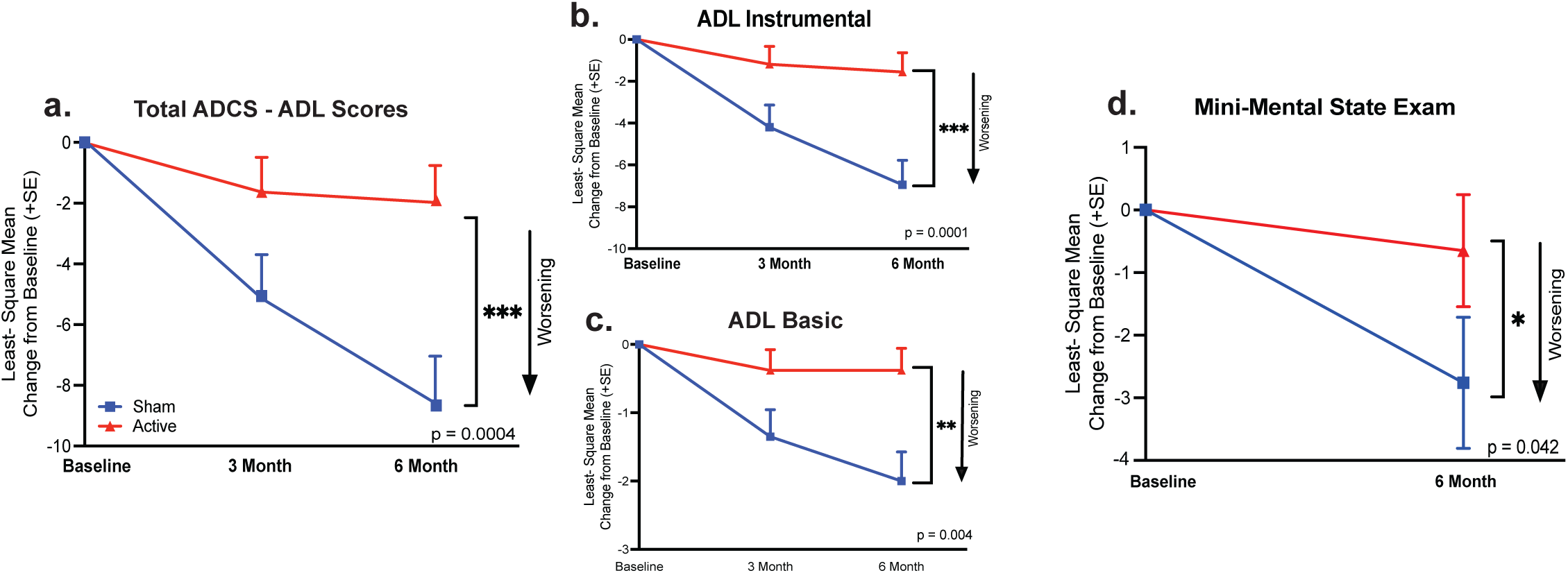
Secondary Outcome Measures. In the Overture trial participants that received active treatment demonstrated slower cognitive decline on the MMSE and ADCS-ADL over the course of 6 months of treatment compared to participants that received sham treatment. Significant differences between active and sham arms at the end of the 6-month trial can be observed across scores on the **a,** ADCS-ADL Total (sham n=26, active n=37; p=0.0004), **b,** Instrumental ADL (sham n=26, active n=43; p=0.0001) and **c,** Basic ADL (sham n=27, active n=43; p=0.004). Additionally, there was a significant difference between the sham (n=27) and active (n=43) groups cognitive trajectories as measured by the **d,** Mini-Mental State Exam (p=0.042). All analyses were conducted using a linear mixed model with significance established as p<0.05. Arrows represent the direction of worsening.

Mini-Mental State Examination (MMSE) scores were evaluated as a secondary outcome, a clinical outcome that is the most broadly used clinical instrument in assessing cognitive function^32^. MMSE scores declined from 19.7±3.85 at baseline to 16.7±6.06 at 6-months in sham arm participants, whereas, decline in the active arm participants was more modest, changing from 21.4±3.28 at baseline to 20.9±4.53 at 6-months. The difference (ΔLSMean ± SE = 2.10±1.07) in decline in MMSE scores between sham and active arm participants was statistically significant (p=0.042; **Figure 3d**). Additional secondary outcome measures included the ADCOMS, Neuropsychiatric Inventory (NPI), Caregiver Burden, Quality of Life (QoL). There were no significant differences between active and sham arms in these instruments (**Extended Data Table 2**). Caregiver Burden, assessed by Zarit Burden Interview (ZBI) scores, showed a 4.6-point increase in sham arm participants, from 21.9±14.81 at baseline to 26.5±19.74 at 6-month, indicting worsening, whereas no changes were observed in the active arm participants (22.8±13.28 at baseline and 24.1±13.81 at 6-month) (**Extended Data Fig. 1**), however, the difference did not reach statistical significance. The Integrated Alzheimer’s Disease Rating Scale (iADRS) was also evaluated, an outcome which combines scores from the ADAS-Cog and ADCS-ADL Instrumental; this outcome can be used in studies of mixed populations of participants from a broad AD disease spectrum^19^. Changes in iADRs scores showed a 11.2-point decline (98.0±20.01 at baseline and 86.8±27.58 at 6-month) in sham group participants and a 3.1-point decline (106.3±14.59 at baseline and 103.2±17.26 at 6-month) in active arm participants, indicating a trend (p=0.09) for a better outcome for active treatment participants (**Extended Data Table 2** & **Extended Data Fig. 1**). In summary, clinical efficacy results indicated that 40Hz sensory stimulation had an effect in clinical measures which are assessing patients broader, more global cognitive and functional abilities and having less or no impact on the measures weighted towards a more narrowly scoped change in cognition, such as ADAS-Cog14.

### MRI BRAIN VOLUME OUTCOMES

Brain atrophy due to neurodegeneration is one of the fundamental pathologies of AD^20^ and brain morphological changes such as whole brain volume loss, cortical thinning, and lateral ventricle expansion have been well documented in both normal aging and in AD by volumetric MRI^21, 22^. In this trial, brain atrophy was evaluated by volumetric MRI measurements (ΔLSMean ± SE) in brain regions closely linked to AD pathology and traditionally monitored in AD trials, such as the whole brain, lateral ventricle, and hippocampus^23, 24^. In sham arm participants, a decline in whole brain volume (– 17.4±3.116) and an (3.55±0.572) increase in lateral ventricle expansion were observed over the 6-month trial period. In active arm participants, a diminished whole brain volume decline (−5.47±2.288 cm3) and a similar expansion (2.93±0.403) in lateral ventricle volume were observed. Although there was no significant change in lateral ventricle expansion between sham and active groups (p=0.3936), there was a highly significant reduction in whole brain volume loss specific to participants that received gamma sensory stimulation, (p=0.005) (**Fig. 4a, b**). In contrast, total hippocampal volume loss was similar between sham (−0.08±0.028 cm3) and active (−0.06±0.019 cm3) arm participants (**Fig 4c**). Since the most profound visually evoked gamma oscillation can be detected in the occipital lobe^25^, changes in response to 40Hz audio/visual stimulation were also studied on occipital lobe volume and occipital cortical thickness. There was a −0.99±0.248 cm^3^ loss in occipital lobe volume and a −0.04 ±0.010 cm^3^ loss in occipital cortical thickness in sham arm participants. Active arm participants had a significantly attenuated loss in occipital volume (difference between arms: −0.18±0.186 cm^3^; p=0.019) and occipital cortical thickness (difference between arms: −0.01±0.007 cm3; p=0.013) compared to sham group participants (**Fig. 4d, e**).

**Figure 4:**
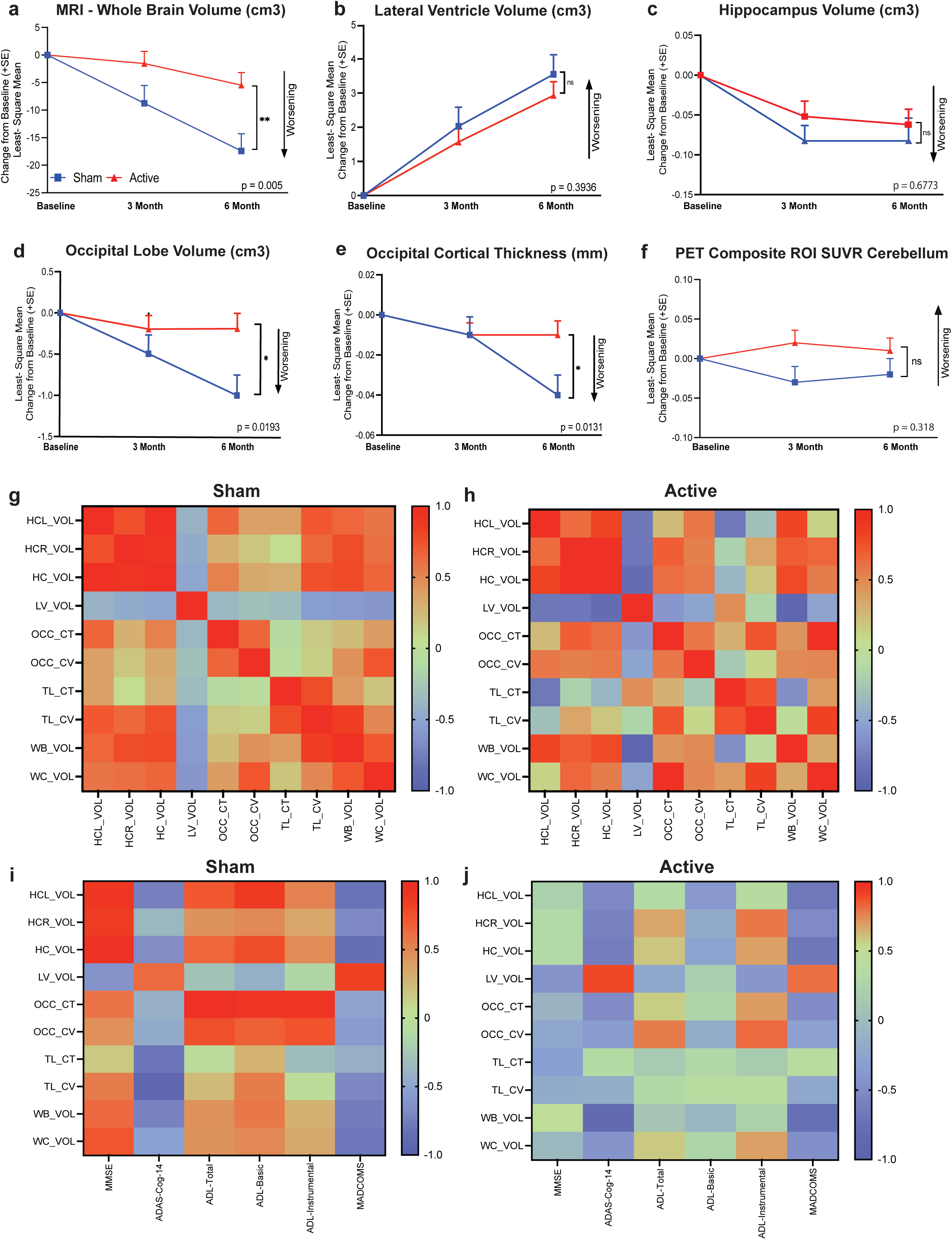
Preservation of brain volume assessed by MRI. Differences in volumetric changes (cm^3^) in **a,** whole brain (sham n=20, vs. active n=36; p=0.005) **b,** lateral ventricle (sham n=19, vs. active n=34; p=0.394), **c,** total hippocampus (sham n=10, vs active n=24; p=0.677), **d,** occipital lobe cm3 (sham n=14, vs. active n=27; p=0.019), and **e,** occipital cortical thickness (sham n=14, vs. Active n=27; p=0.013) as assessed by MRI at baseline and at 3- and 6-months of the therapy. **f,** Composite amyloid PET SUVR values referenced to the cerebellum; no significant difference between active (n=39) and sham (n=23) groups (p=0.318). **g,** Correlation matrix shows R-values for 10 MRI measurements for sham arm (left panel) and **h,** active arm (right panel) participants between various brain regions. (HCL_VOL: Left Hippocampus Volume; HCR_VOL: Right Hippocampus Volume; HC_VOL: Sum of the HCL and HCR Volume; LV_VOL: Lateral Ventricle Volume; OCC_CT Occipital Cortex Thickness; OCC_CV: Occipital cortical volume; TL_CT: Temporal Cortical Thickness; TL_CV: Temporal Cortical Volume; WB_VOL: Whole Brain Volume; WC_Vol: Whole Cortex Volume). Correlation matrixes showing R-values between anatomical changes and clinical outcome measures, including MADCOMS, ADAS-COG-14, ADL Total, ADL Basic, ADL Instrumental, and MMS in **i,** sham arm (left panel) and **j,** active arm (right panel) participants. Correlations shown are r values that were corrected for baseline intercranial volume.

Brain morphological changes were further analyzed by exploring correlations between distinct brain regions, including a previously established inverse correlation between whole brain volume loss and lateral ventricle expansion. In active arm participants, a significant (p=0.0001) inverse correlation was found in volumetric changes between whole brain and lateral ventricles (**Fig. 4g, h**). Additional correlations between whole brain volume and various brain subregions, including the occipital lobe, were also investigated. Summary results of this analysis are shown in **Fig. 4g, h** as correlation coefficient (*r)* values of both active and sham arm groups. Correlations were established between volumetric changes in brain regions that are known to be connected to AD disease progression^21^, including whole brain and temporal cortical lobe volumes (*r*=0.85, p<0.01), whole brain and hippocampus (*r*=0.79, p<0.02), whole brain volume and whole brain cortical volume (*r*=0.72, p<0.04) in sham arm participants. In the active group, the strongest correlations were observed between volumetric changes of whole brain and lateral ventricle (*r*=-0.94, p<0.0001), lateral ventricle and hippocampus (*r*=-0.88, p<0.002), occipital cortical thickness and whole brain cortical volume (*r*=0.97, p<0.00002). In addition, a correlation was found between changes in temporal lobe volume and temporal lobe cortical thickness in both sham (*r*=0.78, p<0.02) and active (*r*=0.74, p<0.02) groups.

Alzheimer’s Disease Neuroimaging Initiative (ADNI) studies have established clear correlations between MRI-based brain atrophy and cognitive and functional abilities in both aging and participants with AD spectrum^26, 27^. Changes in MMSE and ADAS-Cog scores have been shown to strongly correlate with rates of whole brain atrophy and lateral ventricle expansion in MCI and AD patients^26^. Furthermore, it has been shown that atrophy rates of hippocampal or whole brain volume are tracking disease progression and can be considered to serve as outcome measures in clinical trials^24^. Therefore, correlations between brain volumetric changes and changes in clinical outcome measures were also investigated. In the sham group, volumetric changes of the hippocampus correlated with MADCOMS scores (*r*=-0.85, p<0.03) and MMSE (*r*=0.94, p<0.005), occipital cortical thickness with ADCS-ADL total (r=0.97, p<0.002) ADCS-ADL basic (r=0.91, p<0.01) and ADCS-ADL instrumental (*r*=0.92, p<0.01), and temporal lobe volume and cortical thickness with ADAS-Cog14 (*r*=-0.94, p<0.006; *r*=-0.81, p<0.05, respectively) (**Fig. 4i**). In active arm participants, volumetric changes of the whole brain correlated with ADAS-Cog14 (*r*=-0.84, p<0.01) and MADCOMS (*r*=-0.77, p<0.02); lateral ventricle with ADAS-Cog14 (*r*=0.90, p<0.002) and MADCOMS (*r*=0.80, p<0.02); and occipital lobe with ADCS-ADL total (*r*=0.78, p<0.02) and ADCS-ADL instrumental (*r*=0.82, p<0.01) (**Fig. 4j**). It is notable that in active arm participants, a correlation was found between clinical outcome scores and whole brain and occipital lobe volumes, brain regions showing significantly reduced volume loss comparing to sham arm participants. ADCS-ADL scores also correlated with changes in occipital lobe volume and cortical thickness, a relationship previously reported in AD patients^28^.

The volumetric findings together with clinical efficacy data indicate that 6-month sensory stimulation led to considerable preservation of functional and some cognitive abilities, demonstrating 77%, 78%, 81%, slowing in ADCS-ADL total, instrumental and basic scores, respectively, and 76% slowing in MMSE scores in addition to a 69% slowing in whole brain volume loss (**Extended Data Fig. 2**).

### AMYLOID PET IMAGING

While amyloid pathology is an established hallmark of AD pathology, targeting amyloid beta plaques as a therapeutic intervention is still under investigation. Here we compared amyloid pathology at the completion of the 6-month trial period in both active and sham groups. Although PET was conducted in each participant, enrollment was based on clinical presentation, and subsequent amyloid PET results showed that 11 out of 43 participants assigned to the active arm and 9 out of 27 participants assigned to the sham arm were not amyloid positive based on 1.12 cutoff composite SUVR value, using the cerebellum as a reference region. There was no significant difference in SUVR values between active (1.3±0.23, n=39) and sham (1.2±0.25, n=23) groups at baseline, or at the completion of the trial (active arm participant: 1.4±0.22, n=27 and sham; 1.3±0.22, n=18) when both amyloid positive and negative participants were taken into account (**Fig. 4f**). Similarly, no difference between SUVR values were seen when only participants having positive amyloid pathology at baseline were compared to the completion of the trial (active arm: baseline: 1.4±0.16, n=29; 6-month: 1.4±0.16, n=23; sham arm: baseline: 1.4±0.20, n=15, 6-month: 1.4±0.15, n=13). Furthermore, there were no significant differences in any individual brain region values between active and sham arm participants (data not shown).

Since no significant effects on amyloid plaque loads were seen in active arm participants, either in amyloid positive and negative participants or exclusively amyloid positive participants, clinical outcome results were analyzed separately in both amyloid positive and negative participants. The main efficacy outcome results from both amyloid positive and negative participants are summarized in **Extended Data Table 3 and Extended Data Fig. 3**. ADCS-ADL Total scores were significantly different between active and sham groups in both the amyloid positive participant population (ΔLSMean <u>±</u> SE = 5.40<u>±</u>1.67, n=31, p=0.001) and in the amyloid negative participant population (ΔLSMean <u>±</u> SE= 12.45<u>±</u>4.28, n=6, p=0.005) (**Extended Data Fig. 3a, b**). Treatment showed a comparable trend in MMSE scores and in brain volumetric outcomes in amyloid positive and negative participants (**Extended Data Fig. 3c, b**).

## DISCUSSION

Results from the OVERTURE study demonstrated that daily one-hour application of 40Hz auditory and visual stimulation over a six-month period is safe and well tolerated in participants with mild to moderate AD. Exploratory efficacy measurements showed significant benefits of treatment on cognitive and functional outcomes, as assessed by MMSE and ADCS-ADL, when comparing active to sham treated participants. Brain atrophy measured with volumetric MRI was attenuated in participants receiving active treatment. However, some conventional clinical instruments traditionally used in AD trials and assessment of amyloid plaque load did not reveal statistically significant differences between active and sham groups.

The main objective of the Overture trial was to demonstrate the treatment’s safety, tolerability, and feasibility. The excellent safety profile of this treatment is in accord with two previous clinical studies: a delayed start, 4- or 8-weeks trial in 10 participants with mild cognitive impairment^29^, and a single-blinded, randomized, placebo-controlled 3-months, pilot trial in 15 participants with mild probable AD dementia^4^ showing that 40Hz sensory stimulation is safe and well tolerated. In contrast, the present study involved the highest number of treated participants (n=70) in a double-blind sham-controlled randomized trial, over the longest treatment period (6 months), and included both mild- and moderate AD participants. In addition, in contrast to most anti-amyloid monoclonal antibody therapies to date^30^, 40 Hz sensory stimulation did not result in MRI findings of ARIA-E or ARIA-H. The most frequent AEs reported were headache and tinnitus, with higher occurrence in the active arm participants. AEs led to the same proportion of early discontinuation of participants in both arms, indicating no significant differences in treatment limiting tolerability over a 6-month period. The current study also demonstrated very high adherence to the treatment in both active and sham arm participants assessed objectively by device usage. These results provide further evidence that 40Hz audio/visual stimulation is safe and well tolerated by patients on the AD spectrum.

The OVERTURE trial included an extensive selection of exploratory efficacy outcomes to evaluate potential clinical benefits of the treatment. Although a recent publication reported improvement in a face-name associative memory test in mild AD participants following three months of 40Hz auditory/visual stimulation^4^, the efficacy of this treatment has not been assessed by clinical instruments traditionally applied in AD clinical trials. There was a significant difference in the change from baseline for MMSE scores between the active arm and sham arm participants, demonstrating a 76% reduction in decline of cognitive function in active arm participants. In contrast, assessment of cognition using the ADAS-Cog14 did not reveal a significant effect of the treatment. Although ADAS-Cog14 and MMSE values show significant correlation, the reasons for the discrepancy between MMSE and ADAS-Cog14 results are unclear. Historically the ADAS-Cog14 has been considered a sensitive efficacy measure of acetylcholinesterase inhibitors^31^, and therefore might be particularly responsive to cognitive function closely related to cholinergic neurotransmission, including hippocampal memory circuits. Whereas MMSE is the most frequently used cognitive screening instrument in clinical practice^32^, additional studies with advanced, sensitive cognitive assessments are needed to explore which cognitive domains may be most impacted by this treatment.

Decline in functional abilities, assessed by ADCS-ADL, were also significantly diminished by 40Hz auditory-visual stimulation. Both basic and instrumental scores were significantly different between active and sham arm participants, showing that active arm participants maintained their basic and instrumental functional abilities to a comparable degree. These findings indicate a broad beneficial effect of the treatment on multiple functions of AD participants, showing an overall 84% reduction in the decline of functional daily activities. The status of functional abilities is weighted heavily by clinicians when making the distinction between MCI and actual dementia in AD, thus denoting its importance to the clinician. However, to the patient, the presence of impaired instrumental and basic activities of daily living are of greatest importance. It is the decline in functional abilities that most heavily impact the individual’s quality of life, require the need for caregiver assistance, and drive the progression of caregiver burden and the timing of placement in residential assisted living. Indeed, it is when cognitive issues impact function that individuals and their family members are prompted to seek evaluation from their health care provider^33^. Although a slowing in worsening as reflected by the CDR-SB scale was observed, it did not reach statistical significance. A broadly used instrument, iADRS, showed a statistical trend also suggesting beneficial effects of the treatment. An increase in Care Partner burden was observed in sham arm participants over the 6-month treatment period, and no increase was seen in active arm participants, although the difference was not statistically significant between groups. In summary, these promising exploratory outcomes suggest clinically meaningful benefits of 40Hz sensory stimulation in mild-moderate AD. Subsequent clinical trials in larger populations of AD participants evaluated over longer periods of time will inform the spectrum of clinical benefits of this treatment, and whether it will be shown to be uniformly effective in the majority of AD patients, or preferentially demonstrate superior outcomes in certain neurophysiological or pathological subgroups of AD.

Enrolment of participants was based on the clinical features of AD. The amyloid status of participants was determined at baseline and potential changes in amyloid burden were additionally assessed by amyloid PET imaging during the treatment. Experimental studies demonstrated that 40Hz sensory stimulation reduced amyloid Aβ_40_ and Aβ_42_ concentrations in homogenized brain tissue and amyloid plaques were assessed immunocytochemically in transgenic mice^7, 13^. Using PET imaging, the present study did not confirm a reduction in amyloid plaques in response to 40Hz sensory stimulation in participants with AD. However, these results do not exclude the possibility that the treatment could impact soluble amyloid species in the brain which cannot be detected by PET signals. Follow up studies, including measurements of CSF biomarkers will contribute to our understanding of how evoked gamma oscillation could differently affect amyloid pathology in experimental studies and in AD patients. Nevertheless, it can be stated that the currently observed clinical benefits of the treatment are not mediated by reduction in amyloid plaque load, and the clinical benefits were equally demonstrated in amyloid negative participants. These findings also indicate a potential treatment option for patients who are diagnosed with AD on clinical presentation without PET confirmation of amyloid positivity, which is a considerable segment of the dementia population^34^.

The most intriguing result of the OVERTURE trial was the observed reduction in brain volume loss in the active group compared to placebo. A significant reduction in whole brain volume loss was demonstrated with a strong inverse correlation with lateral ventricle enlargement. The present study did not demonstrate reduced hippocampal volume loss, which is contrary to a recent publication demonstrating that three-month, daily, one-hour, 40Hz audio/visual stimulation significantly reduced hippocampal volume loss, assessed by using 3Tesla MRI^4^. In the current study a large proportion of hippocampal scans failed quality control, due to the relatively small hippocampal size and the use of low-resolution 1.5Tesla MRI. However, analysis of the occipital lobe, where the most profound visually evoked gamma oscillation can be detected^25^, a significant reduction in loss of occipital lobe volume and cortical thickness were demonstrated. The current MRI findings of reduced brain volume loss are in accord with experimental results showing that 40Hz sensory stimulation reduces neurodegeneration and brain atrophy via upregulation of cytoprotective proteins and a reduction of DNA damage^9^. Prevention of cortical thinning in the occipital lobe could be due to several mechanisms revealed previously in experimental studies. Repeated application of 40Hz sensory stimulation over a couple of weeks prevented depletion of presynaptic and postsynaptic markers, promoted dendritic spine maturation, and reduced neuronal loss in transgenic mice^9^. The current structural MRI findings indicate that 40 Hz sensory oscillation in mild-moderate AD participants reduces brain atrophy and these volumetric changes are significantly correlated with clinical efficacy outcomes such as ADAS-Cog14 and ADCS-ADL.

The promising results from the OVERTURE trial support the initiation of a randomized, sham-controlled pivotal clinical trial to further explore potential clinical benefits of 40 Hz sensory stimulation in mild-moderate AD patients. This proposed trial will be paired with advanced neuroimaging and established and exploratory AD biomarkers to provide further insight to the mode of action of 40 Hz sensory stimulation in patients. Considering that the treatment has a remarkably benign side-effect profile, together with promising clinical efficacy benefits, this treatment modality could emerge as a safe stand-alone AD treatment or be considered in combination with current or future pharmacotherapies.

## METHODS

### Clinical study participants

The trial enrolled participants of age 50 and older and meeting the National Institute on Aging–Alzheimer’s Association core clinical criteria for probable AD with MMSE scores between 14-26 as diagnosis of mild-to-moderate Alzheimer’s disease^35^. Participants also underwent MRI to exclude those with confounding pathology, such as ischemic stroke, intracerebral macro-hemorrhages or more than 4 micro-hemorrhages, or any findings that prevent volumetric measurements necessary for MRI and PET analysis. Additional exclusion criteria included profound hearing or visual impairment, seizure in medical history, and anti-seizure/anti-epileptic treatment. Stable use of cholinesterase inhibitors was permitted on a stable dose during the trial, however use of memantine was not allowed. Neurophysiological response of participants to sensory stimulation was also assessed, and only participants showing 40Hz steady-state oscillation in response to combined auditory-visual stimulation were included (see below). A reliable lead care partner was also required for enrollment. Main inclusion and exclusion criteria listed in **Extended Data Table 1**. The trial was conducted over 5 clinical sites, including Boston Center for Memory, Netwon, MA (PIs: Paul Solomon, M.D. and Elizabeth Vassey, Ph.D.) Brain Matters Research, Stuart, FL and Del Ray, FL, (PI: Mark Brody M.D.), The Cognitive and Research Center of New Jersey, Springfield, NJ (PI: Michelle Papka Ph.D.), and ActivMed Practices and Research, Methuen, MA (PI: Michael McCartney M.D.)

### Optimization of sensory stimulation for safety and evoked gamma oscillation

Sensory stimulation was carried out with Cognito Therapeutics, Inc. sensory stimulation device. This medical device consists of an eye-set for visual stimulation, headphones for auditory stimulation and a handheld controller. During a clinical visit, the sensory stimulation device was optimized for each participant’s tolerability to the stimuli as well as the presence of measurable 40Hz steady-state oscillation evoked by the auditory and visual sensory stimuli (**Extended Data Fig. 4**). Neural response at 40 Hz during presentation of auditory or visual gamma sensory stimulation was characterized from electroencephalogram (EEG) recordings of neural activity at several different volumes and intensities, respectively. Each stimulus level was presented continuously for 1-4 minutes. All EEG recordings were collected using a 64- or 32-channel ANT-Neuro eego systems [ANT Neuro b.v., Hengelo, Netherlands]. Data were bandpass filtered and re-referenced to the common average of all EEG channels. Power spectral density (PSD) analysis of the responses to each stimulus level was used to estimate 40 Hz neural response. All EEG data analysis was conducted using MATLAB (The Mathworks, Inc. Natick, MA). Following the clinical visit for optimizing sensory stimulation, each participant received their personalized Cognito Therapeutics, Inc. sensory stimulation device. During the therapy, participants could adjust stimulation intensities via the controller within a pre-set range of intensities.

### Clinical study design

This was a multicenter, randomized 6-month, sham-controlled, double blinded clinical trial which took place in the United States (ClinicalTrials.gov identifier: NCT03556280). Participants were assigned at a 2-to-1 ratio into active and sham arms. Participants in both arms used the same medical device and they followed the same treatment protocol, however setting of stimulation parameters of the device was different between arms. Device was preset to evoke 40 Hz steady-state oscillation in active arm participants, whereas in sham arm participants preset stimulation did not evoke 40 Hz steady-state oscillation. Participants, study partners, and assessment raters were blinded to group assignment. The therapy was self-administered at home with the help of a study partner. Participants were required to use the device daily for an hour each day; selection of the 1-hour treatment duration and treatment protocol were based on preclinical findings. Participants were advised to have the treatment in the morning hours, sitting comfortably, and to refrain from unnecessary movement during the stimulation or from falling asleep. The device captured information on day, time and duration of usage, together with level of sensory-stimulation intensities; data was uploaded to a secured cloud server for remote monitoring. The study was conducted in accordance with the Declaration of Helsinki and had ethics committee approval at each participating site. The CIP was developed in accordance with the requirements set forth in the United States Code of Federal Regulation, 21 CFR 812 Investigational Device Exemptions, ISO 14155:2011 Clinical Investigations of Medical Devices for Human Subjects, the Medical Device Directive 93/42/EEC of the European Union, and the Declaration of Helsinki by the World Health Organization (as amended in 2008). Advarra’s IRB Organization (IORG) Number is 0000635 and IRB Registration number is 00000971. All participants provided written informed consent that is adheres to all necessary policies.

### Neuroimaging methods

Magnetic resonance imaging (MRI): All participants in the trial had baseline MRI exams and findings to be in accord with inclusion/exclusion criteria (**Extended Data Table 1**). Acquisition of 1.5 Tesla MRI data at each imaging site followed a previously described standardized protocol^36^ that was rigorously validated across different neuroimaging sites. MRI data was collected for safety consideration and potential impact of the treatment on structural and volumetric changes in the brain. Analysis of MRI scans were assessed for ARIA over the course of the study for safety by a neuroradiologist blinded at treatment arms at a central reading facility (Imaging Corp). All 3D T1-weighted structural MRI images were processed by Biospective, Inc. (Montréal, QC H3B 2T9, Canada) using their proprietary PIANO™ software. PIANO™ is a configurable, modular, pipeline-based system for fully automated processing of multi-modality images. PIANO™ was designed for high-throughput processing of large-scale, multi-center, neuroimaging data. PIANO™ was configured and validated specifically for this study to incorporate the study-specific details utilized by the core PIANO™ modules. For the present study, a standard ADNI template and corresponding atlas was used. The regions of interest (ROIs) were defined on this anatomical template and included whole brain volume (cerebrum and cerebellum), including only brain parenchyma (not CSF), whole brain cortex (cerebral cortical gray matter), lateral ventricle, occipital and temporal lobe volumes and left and right hippocampal volumes. Cortical thickness was computed at each vertex as the distance between the inner and outer surfaces^37^. Parametric maps were mapped to a standardized surface by non-rigid 2D surface registration in order to derive the spatially normalized measures^38^. Cortical thickness values were generated for the following ROIs: composite temporal lobe (entorhinal, fusiform, inferior temporal gyrus, middle temporal regions) and occipital lobe (lingual, cuneus, superior occipital gyrus, inferior occipital gyrus, calcarine sulcus, middle occipital gyrus). All volumetric MRI analyses were controlled for total intracranial volume.

### Amyloid-β Positron Emission Tomography (PET) Imaging

Amyloid pathology was determined during enrolment, 3 and 6 months of treatment using Amyvid [18F] florbetapir positron emission tomography (PET) imaging, performed 50-70 minutes following the intravenous administration of the tracer. PET images were obtained from the level of the vertex to the base of the skull. Axial, sagittal, and coronal views were provided and qualitatively classified as amyloid positive or negative based on the Amyvid prescribing pharmaceutical guidelines. For quantitative analysis, the amyloid PET images underwent several processing steps using Biospective’s PIANO™ software, including frame-to-frame motion correction, image smoothing, and co-registration to anatomical MRI. Following linear registration to the participant-specific, baseline T1-weighted, 3D anatomical MRI volume, the PET volumes were spatially normalized to reference space using the linear transformations derived from the anatomical MRI registration. ROI-based standardized uptake value ratio (SUVR) measures, generated by PIANO™, were obtained using automated atlas-based parcellation in stereotaxic space using the whole cerebellum and pons as reference regions. Composite ROI (frontal cortex, lateral temporal cortex, parietal cortex, somatosensory cortex, cuneus) values are presented here.

### Safety and Tolerability Endpoints

The primary objective was to evaluate the safety and tolerability. Safety was assessed as the incidence and nature of adverse events (AEs) based on the safety population. AEs were coded using the Medical Dictionary for Regulatory Activities (MedDRA v16.1). Severity levels include mild, moderate and severe. Therapy relationships were grouped into two categories: related and unrelated. Unrelated and unlikely were categorized as “unrelated.” Possible, probable and definite were categorized as “related.” If a participant has the same AE on multiple occasions, the highest severity or therapy relationship recorded for the event will be presented. The tolerability endpoint was adherence to treatment regime as measured by daily device use.

As a Phase1/Phase2, exploratory trial, Overture utilized a broad range of efficacy outcome measures selected based on their previous applications in AD clinical trials. However, without *a priori* understanding of potential clinical benefits in patients all efficacy outcome measures are considered exploratory in this trial. The trial was not statistically powered to evaluate potential efficacy outcomes, nor were the previous clinical trials that evaluated 40 Hz gamma sensory stimulation in participants with mild to moderate AD. Clinical instruments assessing functional and cognitive outcomes included: Alzheimer’s Disease Cooperative Study Activities of Daily Living Inventory (ADCS-ADL), CRD, ADAS-Cog14, and MMSE. AD Composite Outcomes scores were also created, such as ADCOMS, (comprising items from the ADAS-Cog 14, MMSE, and CDR-SB) to index cognition and function^39^. MADCOMS (optimized to mild and moderate patients, consisting of 7 items, 3 ADAS–Cog subscale items, 2 MMSE items, and 3 CDR SB items) and Integrated Alzheimer’s Disease Rating Scale (iADRS); unweighted composite of the ADAS-Cog14 (all items) and the Instrumental subscale of the ADCS-ADL, reflecting relative equal contributions from cognitive and functional domains^19^. Additional clinical instruments included Neuropsychiatric Inventory (NPI), Caregiver Burden (Zarit Burden Interview ZBI), Columbia–Suicide Severity Rating Scale and Quality of Life (QoL).

### Statistical Analysis

Efficacy endpoints were analyzed in the intent to treat population based on treatment administered (ITT; received at least one day of therapy and one post-baseline assessment) by comparing the change in outcomes between initial assessment and 26-weeks between treatment groups using a linear mixed model with repeated measures. This model used all available data. Missing data were not imputed except to generate the MMSE month 3 data (straight-line imputation from initial assessment to 6 months) for use in the MADCOMs composite metric. The model assessed whether or not there was a difference in estimated change from initial assessment values between treatment group and sham at 26 weeks. Treatment group and time were included as a fixed effect. Clinical site and intercept were included as random effects. Age, baseline MMSE, and baseline efficacy parameter were used as covariates and treatment by time and treatment by baseline interactions were included in the model. Intercranial volume was used as a covariate in all volumetric and cortical thickness analyses.

## Data Availability

The data that support the findings of this study are available on request from the corresponding author [M.H.]. The data are not publicly available due to them containing information that could compromise research participant privacy/consent.

## Author Contributions

Z.M., J.T.M., E.H., K.K. designed the clinical study. J.N-J., S.H. were clinical study statisticians. M.H., A.B., E.H., M.S., A.K., C.V.S., V.F., B.V., Z.M., J.T.M. analyzed and/or interpreted clinical study data. M.H., A.B., M.S., J.T.M., Z.M. wrote the manuscript. All authors approved the final version of the manuscript for submission.

## ACKNOWLEDGEMENTS

Authors are grateful to clinical trial participants and their care partners, and clinical team members, including clinical site principal investigators Paul Solomon, M.D. and Elizabeth Vassey, Ph.D. (Boston Center for Memory, Netwon, MA, Mark Brody M.D. (Brain Matters Research, Stuart, FL and Del Ray, FL), Michelle Papka Ph.D. (The Cognitive and Research Center of New Jersey, Springfield, NJ), and Michael McCartney M.D. (ActivMed Practices and Research, Methuen, MA).

---

**Extended Data Figure 1:**
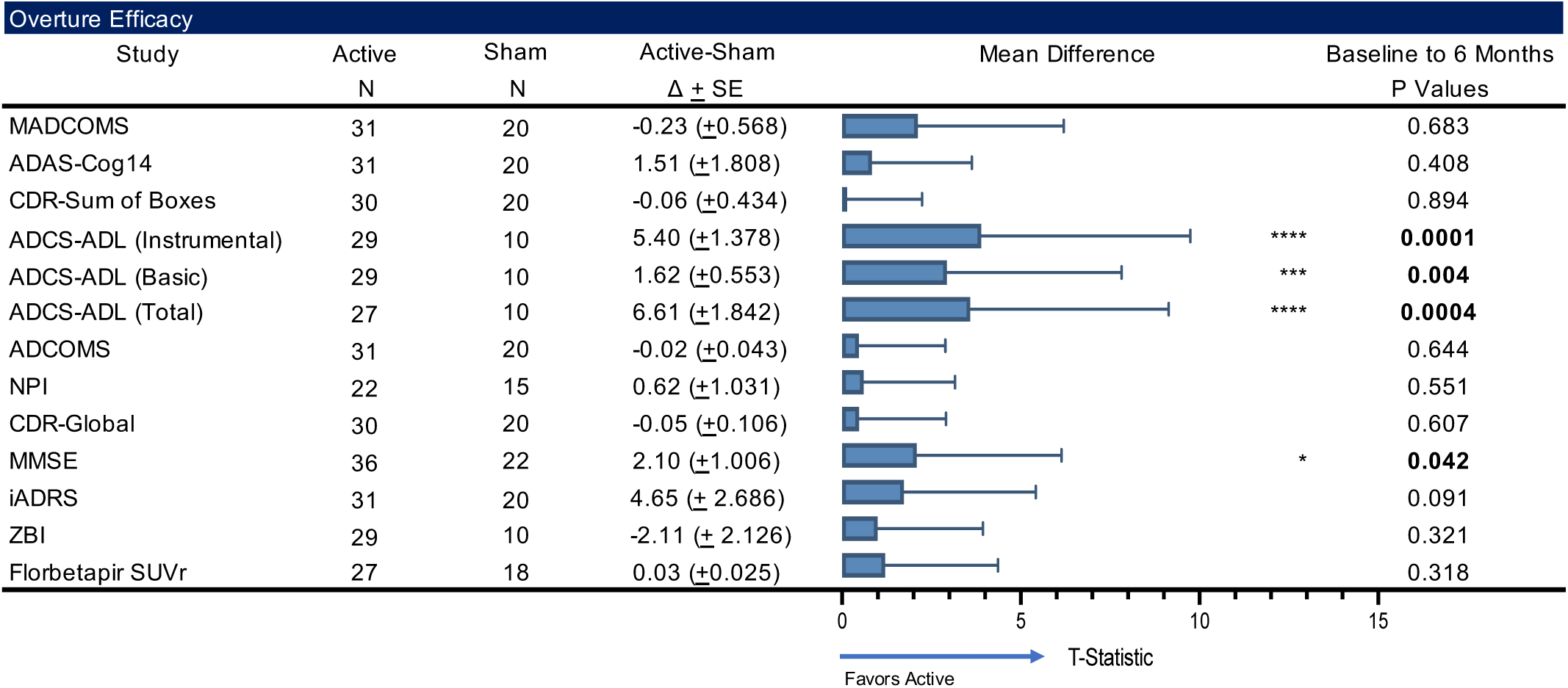
Forest plot of overture efficacy outcomes. Forest plot of LSMeans(SD) of efficacy outcomes to explore the association between the sham and active arms for each subgroup. Statistical values are represented in the table (N, LSMeans, and P Values).

**Extended Data Figure 2:**
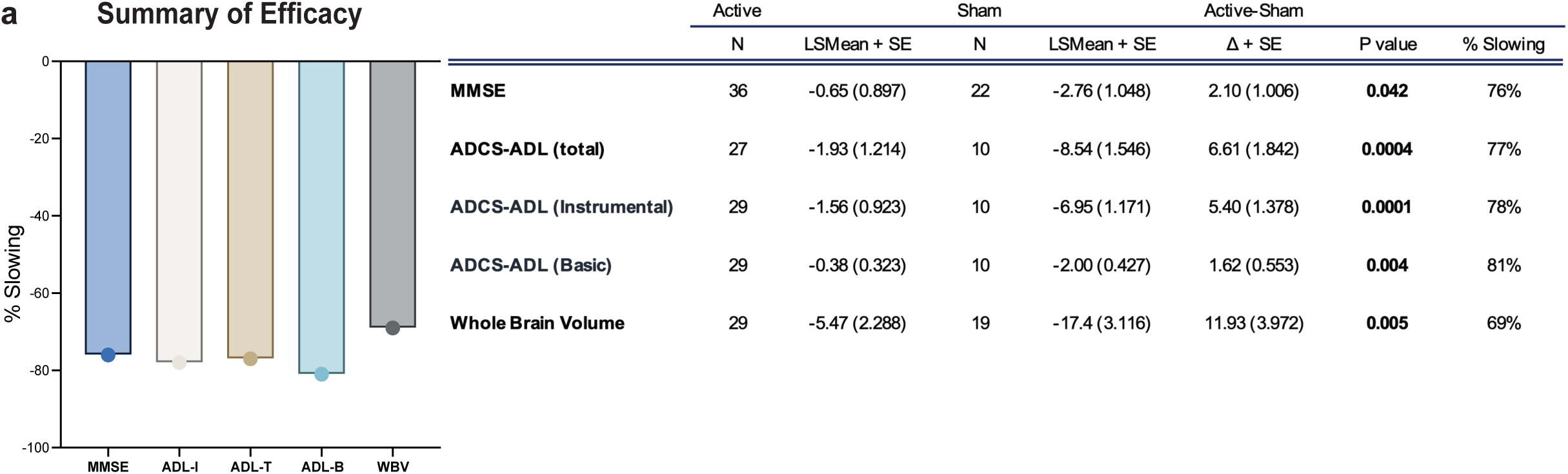
Summary of efficacy data presents the percent slowing, **a**, calculated as the difference in means for active vs sham, for functional outcomes measures (MMSE (76%), ADL (total (77%), basic (78%), instrumental (81%), and whole brain volume (69%).

**Extended Data Figure 3:**
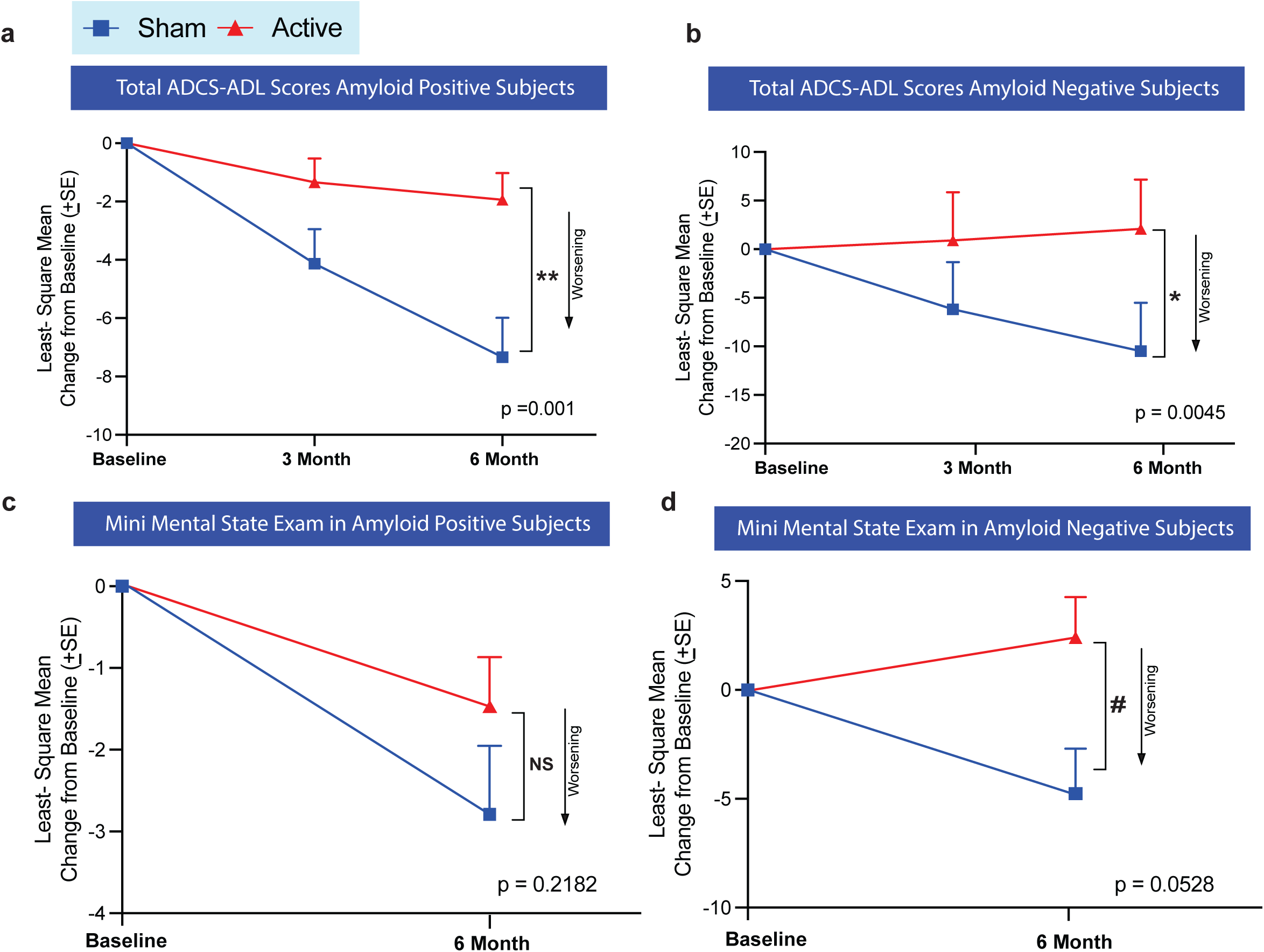
40Hz steady state oscillation therapy is effective in both amyloid positive and amyloid negative participant. **a**, ADCS-ADL total score change in amyloid positive participants in sham (n=8) vs active (n=23) participants at the end of the 6-month trial (p=0.001). b, ADCS-ADL total score change in amyloid negative participants in sham (n=2) vs active (n=4) at the end of the 6-month trial (p=0.005). **c**, MMSE total score change in amyloid positive participants in sham (n=15) vs active (n=27) at the end of the 6-month trial (p=0.218). **d**, MMSE total score change in amyloid negative participants in sham (n=7) vs active (n=9) at the end of the 6-month trial (0.053). All analysis was conducted using a linear mixed model with significance established as p<0.05.

**Extended Data Figure 4:**
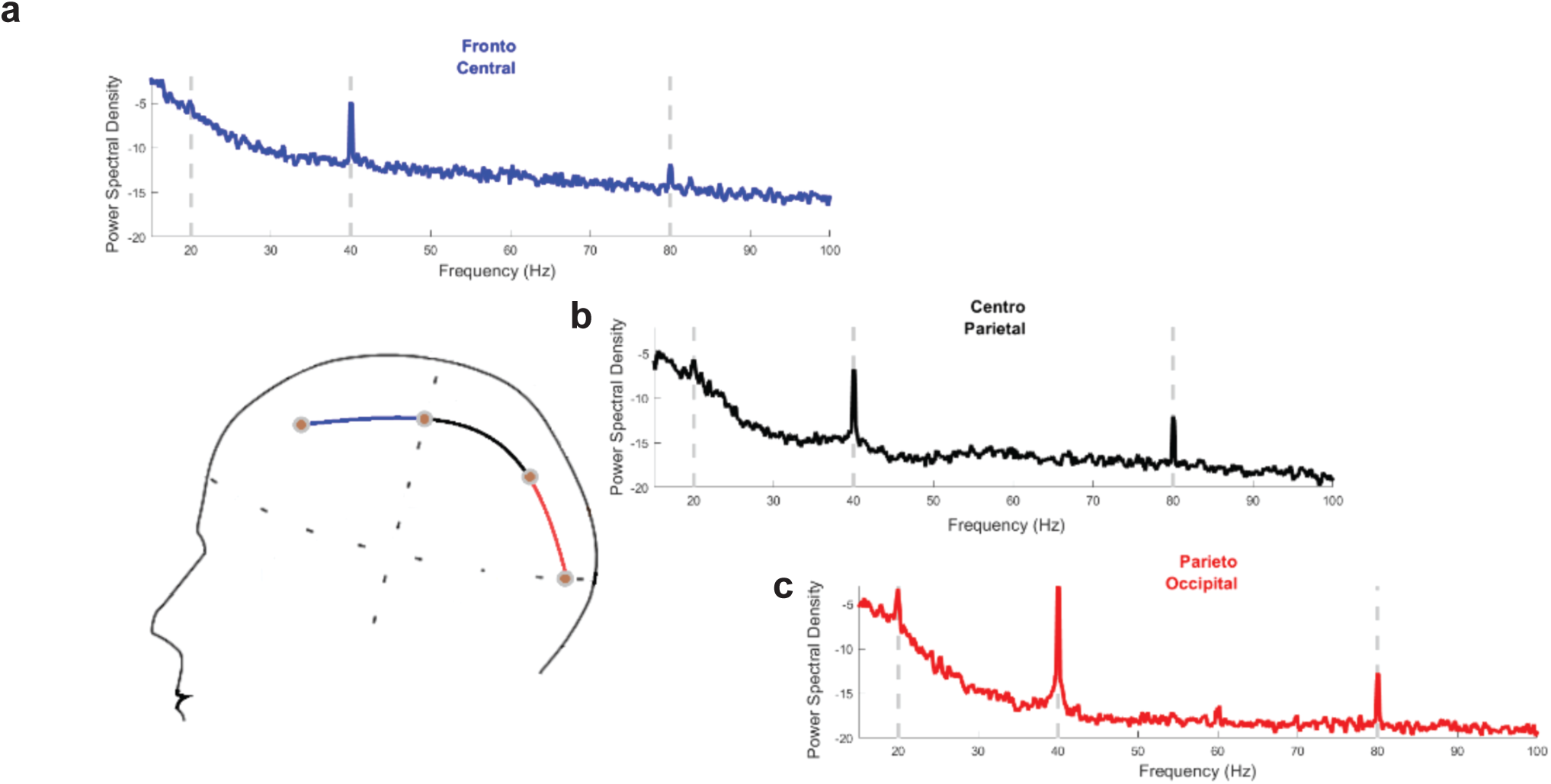
Neuronal activity as observed from electroencephalogram (EEG) during 40Hz audio and visual gamma sensory stimulation from an individual with Alzheimer’s Disease. Neural response at 40Hz along with subharmonics and superharmonics of 40Hz can be observed in large cortical regions. Shown here are the neural activity from **a**, Fronto-Central, b, Centro-Parietal, and **c**, Parieto-Occipital regions. In the **c**, Parieto-Occipital region the observed response is more pronounced.

**Extended Data Table 1:** Inclusion and Exclusion Criteria

| Inclusion Criteria |  |
| --- | --- |
| Subjects who met the following criteria were considered eligible to participate in the study: |  |
| 1 | Was $\geq 50$ years old at the time of consent. |
| 2 | Had a MMSE score ranging from 14-26, inclusive. |
| 3 | Had a diagnosis of a clinical syndrome of cognitive impairment consistent with prodromal AD, AD, or MCI due to AD per National Institute on Aging-Alzheimer's Association (NIA-AA) diagnostic criteria. |
| 4 | Could identify (or had already identified) a legally authorized representative who can verify study inclusion/exclusion criteria. |
| 5 | Had a reliable Lead Care Partner or informant (defined as an individual who knew subject well and had contact with subject for at least 10 hours each week or in investigator's opinion had sufficient knowledge of the subject's daily life to be a reliable informant and attend to the subject during treatment sessions) who also consented to study participation. |
|  | NOTE: Up to 2 Alternate Care Partner(s) could be designated as well as a Lead Care Partner, provided each person also attended a clinic visit and signed the ICF and attended Initial Therapy or First At Home Therapy for training on device use from Field Support. |
| 6 | Showed adequate induced activity (entrainment) during baseline EEG. |
| 7 | Provided consent or assent (as appropriate) to participation in study assessments. |

| Exclusion Criteria |  |
| --- | --- |
| Subjects who met any of the following criteria were not included in the study: |  |
| 1 | Self- or caregiver reported concurrent profound hearing or visual impairment. |
| 2 | Self- or caregiver reported recent or clinically relevant history of seizure. |
| 3 | Active treatment or concurrent prescription for the purpose of preventing seizure with one or more anti-seizure/anti-epileptic. |
| 4 | Prior ischemic stroke, intracerebral hemorrhage, or subarachnoid bleed within the past 24 months, or that caused MRI exclusion criteria to be met. |
| 5 | Self- or caregiver reported usage of any new AD medication within the past 30 days, or concurrent/expected titration of dosage of any AD medications during the study period. |
| 6 | NOTE: AD medications and dosage had to be stable for 30 days at baseline neuropsychological assessments and initial therapy date. If doses were changed during screening, neuropsychological assessments had to be repeated. |
| 7 | Active treatment or concurrent prescription with memantine (Namenda™, Namzaric™) within 30 days of baseline neuropsychological assessments and starting therapy. |
| 8 | Active treatment or concurrent prescription with medications that lower seizure threshold and pose a risk for participation in the study investigator's opinion. |
| 9 | Self- or caregiver reported of physician-diagnosis of a medical, neurological/neurodegenerative or psychiatric condition that in the opinion of the investigator could be contributing to the patient's cognitive impairment. This could include, but was not limited to, vitamin B12 deficiency, abnormal thyroid function, stroke or other cerebrovascular condition, traumatic brain injury, Lewy Body disease, Parkinson's disease, fronto-temporal dementia, schizophrenia, bipolar disorder, or uncontrolled major depressive disorder. Self- or caregiver report of current or historic migraines that in the investigator's opinion would potentially interfere with study compliance. |
| 10 | Self- or caregiver reported of physician-diagnosis of recent or clinically relevant major depressive disorder. |
| 11 | Self-report of clinically significant psychiatric illness or behavioral problem or use of psychiatric agent that could interfere with study completion, as determined by study physician. |
| 12 | Self- or caregiver reported alcohol or substance abuse within the past year. |
| 13 | Self- or caregiver reported concurrent enrollment in any anti-amyloid clinical trial within 30 days or 5 half-lives of the investigational intervention (whichever was longer) of the baseline neuropsychological assessments, baseline PET and starting therapy. |
| 14 | Subjects who, in the investigator's opinion would not comply with study procedures. |
| 15 | Subjects with active implantable neurological devices including deep brain stimulators (DBS) and/or non-MRI-safe implantable devices including pacemakers, implantable cardioversion defibrillators (ICDs), spinal cord stimulators, and non-MR compatible surgical implants. |
| 16 | Subject was pregnant, lactating, or of childbearing potential (i.e. women had to be 2 years post-menopausal, used 2 forms of birth control, or surgically sterile). |
| 17 | Previous amyloid imaging with 18F-AV-45 within the previous 3 months of baseline PET scan. |
| 18 | Concurrent or recent participation in any procedures involving radioactive agents such that the total radiation dose exposure to the subject in any given year would exceed the limits of annual and total dose commitment set forth in the US CFR Title 21 Section 361.1. |
|  | Any of the following MRI findings: |
|  | a. Prior macro-hemorrhages |
|  | b. More than 4 micro-hemorrhages |
|  | c. Any findings that prevent volumetric measurements necessary for PET analysis |
| 19 | Investigator opinion that it was not in the best interest of the subject to participate in the study. |

**Extended Data Table 2:** Additional ITT As Treated Efficacy Measures.

|  | ITT As Treated - Efficacy Measures |  |  |  |  |  |
| --- | --- | --- | --- | --- | --- | --- |
|  | Active |  | Sham |  | Active-Sham |  |
| | N | Mean ± SE | N | Mean ± SE | $\Delta \pm$ SE | P value |
| ADCOMS | 31 | 0.13 (0.034) | 20 | 0.15 (0.041) | -0.02 (0.043) | 0.644 |
| NPI | 22 | 0.09 (0.810) | 15 | -0.53 (0.964) | 0.62 (1.031) | 0.551 |
| ADAS-Cog 14 | 31 | 3.55 (1.416) | 20 | 2.05 (1.745) | 1.51 (1.808) | 0.408 |
| iADRS | 31 | -5.19 (1.486) | 20 | -9.83 (2.098) | 4.65 (2.686) | 0.0906 |
| CDR-Global | 30 | 0.18 (0.056) | 20 | 0.24 (0.080) | -0.05 (0.106) | 0.607 |
| CDR-Sum of Boxes | 30 | 1.21 (0.298) | 20 | 1.27 (0.381) | -0.06 (0.434) | 0.894 |
| Care Partner Burden | 29 | -0.15 (1.835) | 10 | 1.96 (2.153) | -2.11 (2.126) | 0.321 |
| QoL-AD (Self Report) | 29 | 0.67 (0.637) | 9 | 0.78 (0.868) | -0.11 (1.115) | 0.919 |
| QoL-AD (Family Report) | 29 | -0.12 (0.578) | 10 | -1.22 (0.775) | 1.10 (1.001) | 0.272 |
| Temporal Cortical Volume | 27 | -1.77 (0.263) | 14 | -1.37 (0.417) | -0.40 (0.510) | 0.441 |
| Temporal Cortical Thickness | 27 | -0.03 (0.009) | 14 | -0.02 (0.015) | -0.01 (0.018) | 0.573 |
| Right Hippocampus | 21 | -0.01 (0.031) | 12 | -0.03 (0.034) | 0.02 (0.028) | 0.568 |
| Left Hippocampus | 22 | -0.03 (0.010) | 15 | -0.04 (0.014) | 0.01 (0.018) | 0.679 |
| Total Hippocampus | 19 | -0.006 (0.019) | 11 | -0.08 (0.028) | 0.01 (0.033) | 0.677 |
| Whole Cortex Volume | 27 | -6.79 (1.671) | 15 | -9.51 (2.509) | 2.71 (3.126) | 0.392 |

**Extended Data Table 3:**
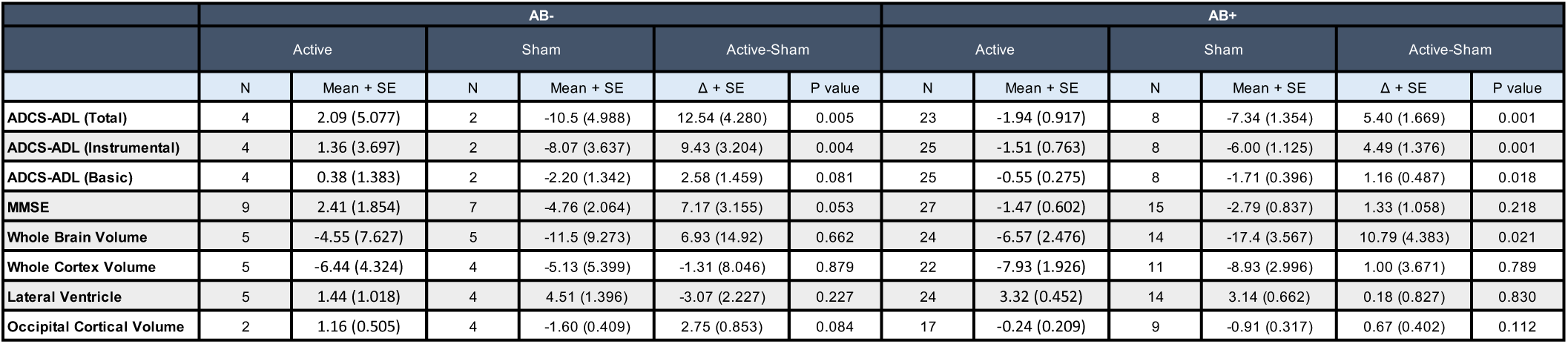
Efficacy measure segmentation between amyloid beta positive and negative participants

